# Predicting COVID-19 related death using the OpenSAFELY platform

**DOI:** 10.1101/2021.02.25.21252433

**Authors:** The OpenSAFELY Collaborative, Elizabeth J Williamson, John Tazare, Krishnan Bhaskaran, Helen I McDonald, Alex J Walker, Laurie Tomlinson, Kevin Wing, Sebastian Bacon, Chris Bates, Helen J Curtis, Harriet Forbes, Caroline Minassian, Caroline E Morton, Emily Nightingale, Amir Mehrkar, Dave Evans, Brian D Nicholson, Dave Leon, Peter Inglesby, Brian MacKenna, Nicholas G Davies, Nicholas J DeVito, Henry Drysdale, Jonathan Cockburn, Will Hulme, Jess Morley, Ian Douglas, Christopher T Rentsch, Rohini Mathur, Angel Wong, Anna Schultze, Richard Croker, John Parry, Frank Hester, Sam Harper, Richard Grieve, David A Harrison, Ewout W. Steyerberg, Rosalind M Eggo, Karla Diaz-Ordaz, Ruth Keogh, Stephen JW Evans, Liam Smeeth, Ben Goldacre

## Abstract

**Objectives:** To compare approaches for obtaining relative and absolute estimates of risk of 28-day COVID-19 mortality for adults in the general population of England in the context of changing levels of circulating infection.

**Design:** Three designs were compared. (A) case-cohort which does not explicitly account for the time-changing prevalence of COVID-19 infection, (B) 28-day landmarking, a series of sequential overlapping sub-studies incorporating time-updating proxy measures of the prevalence of infection, and (C) daily landmarking. Regression models were fitted to predict 28-day COVID-19 mortality.

**Setting:** Working on behalf of NHS England, we used clinical data from adult patients from all regions of England held in the TPP SystmOne electronic health record system, linked to Office for National Statistics (ONS) mortality data, using the OpenSAFELY platform.

**Participants:** Eligible participants were adults aged 18 or over, registered at a general practice using TPP software on 1^st^ March 2020 with recorded sex, postcode and ethnicity. 11,972,947 individuals were included, and 7,999 participants experienced a COVID-19 related death. The study period lasted 100 days, ending 8^th^ June 2020.

**Predictors:** A range of demographic characteristics and comorbidities were used as potential predictors. Local infection prevalence was estimated with three proxies: modelled based on local prevalence and other key factors; rate of A&E COVID-19 related attendances; and rate of suspected COVID-19 cases in primary care.

**Main outcome measures:** COVID-19 related death.

**Results:** All models discriminated well between patients who did and did not experience COVID-19 related death, with C-statistics ranging from 0.92-0.94. Accurate estimates of absolute risk required data on local infection prevalence, with modelled estimates providing the best performance.

**Conclusions:** Reliable estimates of absolute risk need to incorporate changing local prevalence of infection. Simple models can provide very good discrimination and may simplify implementation of risk prediction tools in practice.

## INTRODUCTION

Within 9 months of being characterised as a pandemic by the World Health Organization^1^, cases of COVID-19 had reached almost 100 million globally and around 3.8 million in the UK, with more than 2 million deaths attributed to the virus globally and 100,000 in the UK.^2,3^ Evolving policies regarding shielding, return-to-work guidance, prioritisation of vaccinations and individual choices about restricting social contact are heavily informed by estimated risk of severe outcomes from COVID-19. Here, we focus on risk prediction in the general population, in contrast to predicting prognosis among hospitalised or test-positive subgroups. Estimates of absolute risk are desirable to inform policy and public health decisions. However, transporting estimates of absolute risk from one context to another, such as a different time period or a different geographical region, is particularly challenging in COVID-19, due to the substantial variation in the prevalence of infection over time and by geography.^4^ Prediction models that do not explicitly model the underlying prevalence of infection are likely to produce poor absolute risk estimates. However, the prevalence of infection is not directly measured thus proxy measures must be used. Whether easily accessible proxy measures are sufficiently good to produce reasonable absolute and relative risk estimates remains uncertain.

Early risk prediction models relating to COVID-19 outcomes were found to be poorly reported and at high risk of bias and over-optimism.^5^ However, two subsequent models have been developed to predict risk scores for COVID-19 death in the UK general population. The COVID-AGE is a risk score obtained by an algorithm derived by combining evidence from published studies.^6,7^ QCOVID is a prediction model estimated using routinely collected primary care data in the UK.^8^ Both of these risk prediction approaches met most criteria for low risk of bias in the development of risk prediction algorithms.^9^ However, neither of these algorithms involved explicit modelling of the underlying prevalence of COVID-19 infection, which limits the extent they could be adapted to provide accurate estimates of absolute risk for time periods or settings with different infection prevalences.

In this study, we therefore used data held in the OpenSAFELY platform^10^ on almost 12 million adults in England to answer: 1) How well can we improve estimates of absolute risk of COVID-19 mortality by explicitly incorporating proxy estimates of the changing infection prevalence? 2) Do risk prediction models which do not explicitly model the underlying burden of infection produce a good ranking of people’s risk of COVID-19 mortality in different contexts (time and geographical)? 3) Can simpler prediction algorithms be used without losing substantial predictive ability, compared with more richly specified models?

## METHODS

Full details of the methods used can be found in our pre-published protocol.^11^ This manuscript follows the TRIPOD statement for prediction models.^12^

### Data Source

Primary care records managed by the GP software provider TPP were linked to ONS death data through OpenSAFELY, a data analytics platform created by our team on behalf of NHS England to address urgent COVID-19 research questions (https://opensafely.org). OpenSAFELY provides a secure software interface allowing the analysis of pseudonymized primary care patient records from England in near real-time within the EHR vendor’s highly secure data centre, avoiding the need for large volumes of potentially disclosive pseudonymized patient data to be transferred off-site. This, in addition to other technical and organisational controls, minimizes any risk of re-identification. Similarly pseudonymized datasets from other data providers are securely provided to the electronic health record vendor and linked to the primary care data. The full dataset within OpenSAFELY is based on 24 million people currently registered with GP surgeries using TPP SystmOne software. It includes pseudonymized data such as coded diagnoses, medications and physiological parameters. No free text data are included.

#### Study Population

The target population of interest is adults in England living in the community; residential settings are excluded since risks experienced in institutions such as care homes are likely to be very different to those in smaller households.

The base cohort used in this study comprises males and females aged 18 years or older registered as of 1^st^ March 2020 in a general practice employing the TPP system. Patients with missing age or a recorded age over 105 years, missing sex or missing postcode (from which household and geographic information is calculated) were excluded. Households of greater than 10 people were excluded. The study timeframe was the 100 day period beginning 1^st^ March 2020 and ending 8^th^ June 2020.

### Study Measures

#### Outcome

The outcome to be predicted is 28-day risk of COVID-19 related death. Risk is predicted for the general community, rather than infected people, thus the risk being predicted combines the risk of infection and the risk of dying once infected. Primary care records held within the TPP system were linked to mortality data from the Office for National Statistics (ONS). COVID-19 related death was defined as a death with an ICD-10 code of U07.1 or U07.2 anywhere on the death certificate.

#### Predictor variables

We selected candidate predictors based on known or plausible associations with exposure to COVID-19 infection, risk of severe illness or respiratory tract infection, and factors associated with healthcare access or level of care. Our data include diagnoses (Read 3 CTV3), prescriptions (dm+d), basic sociodemographics and vital signs. Briefly, potential predictors included: age, sex, ethnicity, deprivation, number in household, presence of young children (under 12 years old) in household, a rural indicator, obesity, smoking and blood pressure. Comorbidities included: diagnosed hypertension, chronic heart disease, atrial fibrillation, surgery for peripheral arterial disease, deep vein thrombosis or pulmonary embolism, diabetes, stroke, dementia, other neurological conditions, asthma, respiratory disease, haematological and non-haematological malignancies, solid organ transplant, dialysis, poor kidney function, common autoimmune diseases, asplenia, other immunosuppressive conditions, inflammatory bowel disease, HIV, learning disability, serious mental illness, and fragility fracture in the last two years. Details and codelists are provided in the Appendix, Table A1.

#### Proxy measures of the prevalence of COVID-19 infection

Three different proxy measures of infection prevalence, measured daily, were considered. First, modelled estimates were obtained from dynamic disease modelling,^13^ with estimates obtained by region (7 regions in England) and by 5-year age-group. These estimates account for the infection prevalence, the way in which different age-groups interact with each other and the proportion of the population who are susceptible. These are estimates, thus come with uncertainty and potential error, neither of which is accounted for within our modelling. Second, the mean daily rate of COVID-19 related A&E attendances over the last 7 days was estimated within each Sustainability and Transformation Partnership (STP; used as a measure of local geographic area). Rate of A&E attendances is likely to be an imperfect proxy since it is likely to lag behind true prevalence of infection. Third, the mean daily rate of suspected COVID-19 cases (with CTV3 Codes XaaNq, Y20cf, Y211b, Y22b7 and Y22b8 indicating a suspected case) in primary care over the last 7 days was estimated by STP. A&E attendances and suspected cases in primary care are both likely to lag behind the true infection prevalence, although this will not necessarily hinder performance in predicting COVID-19 mortality. They may also be sensitive to changes in how and when people interact with primary care providers.

### Study design

Three approaches were used: (A) a case-cohort study, (B) 28-day landmarking,^14^ and (C) daily landmarking. The first is a computationally efficient version of a traditional cohort design which does not explicitly model changes in the infection prevalence. The second involves creating multiple sequential overlapping 28-day sub-studies, allowing time-varying proxy measures of the infection prevalence to be explicitly incorporated into the risk prediction model. The third approach additionally updates the measures of the infection prevalence throughout the 28-day period, to try to better estimate the relationship between current infection prevalence and risk.

For approach A, follow-up began 1^st^ March 2020 and ended at the first of: COVID-19 related death or 8^th^ June 2020. The outcome was COVID-19 related death. No censoring was applied at death due to non-COVID causes, because the target of inference was the sub-distribution hazard. The sub-distribution hazard can be estimated by simply not censoring participants at the competing event because our only censoring event is the competing event of death due to other causes.^15^ The analysis sample included all cases of COVID-19 related death and a random age-stratified sample of the eligible patient population (the ‘sub-cohort’, largely comprising non-cases but likely to contain some cases by chance),^16,17^ with sampling fractions of 0.01 in the age-group 18-<40, 0.02 in 40-<60, 0.025 in 60-<70, 0.05 in 70-<80 and 0.13 in 80+ years.

For approach B, a series of 73 overlapping sequential sub-studies were extracted from the base cohort. The sub-studies started 0, 1, 2, 3, 4…, 72 days after 1^st^ March 2020 and each sub-study continued for exactly 28 days. The first sub-study began on 1^st^ March and the last began on 12^th^ May 2020 (Figure 1). All patients from the base cohort still alive at sub-study start were eligible to participate in that sub-study. Follow-up started at sub-study entry date and ended at the first of COVID-19 related death or 28 days after sub-study entry. Participants were not censored at deaths due to other causes. The outcome was COVID-19 related death during the 28 day period. Each sub-study had a case-cohort design, including all eligible patients who experienced a COVID-19 related death during the 28-day sub-study period as cases and an age-stratified random sample of sub-study eligible patients (the sub-study sub-cohort), with age-group specific sampling fractions equal to 1/70 of the sampling fractions for approach A (e.g. 0.01/70 = 0.00014 in the age-group 18-<40). Data from the 73 sub-studies were combined for analysis. Predictor variables and proxy measures of the infection prevalence were assessed at day 0 of each sub-study.

**Figure 1.**
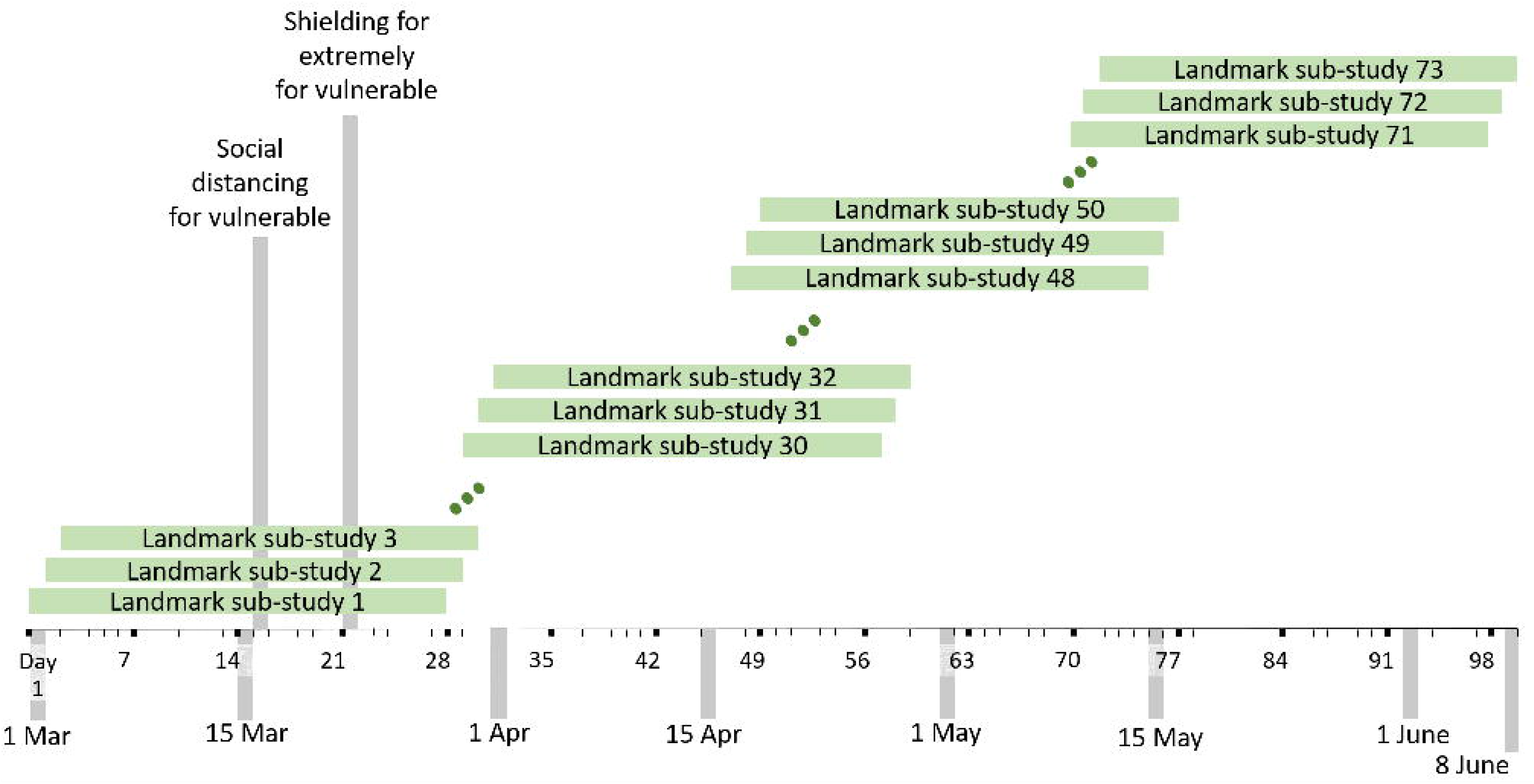
Schematic showing the design of the 28-day landmarking sub-studies (approach B)

Approach C also used a series of stacked sub-studies, with each lasting a single day. Thus 100 sub-studies were formed, the first starting on 1^st^ March 2020 and the last starting on 8^th^ June 2020. Each sub-study included all cases (COVID-19 related deaths) occurring on that day and a random age-stratified sample of non-cases who remained alive by the previous day, with sampling fractions equal to 1/100 of the sampling fractions for approach A. The outcome was the binary outcome of whether or not the sub-study participant experienced a COVID-19 related death on that day. This approach also required information about the daily rate of death due to other causes, which was estimated in a second case-cohort sample, comprising a sampling fraction of 0.3 of all non-COVID-19 related deaths on each day and an age-stratified sample of participants who did not die of non-COVID-19 related causes on that day, with sampling fractions equal to 1/100 of the sampling fractions for approach A.

### Statistical analysis

Variable selection was undertaken from the set of candidate predictors using a penalised regression approach (lasso)^18^. Functional forms for the proxy measures of burden of disease were selected using Akaike’s Information Criterion, as detailed in the protocol.^11^

For approach A, a Cox proportional hazards model was fitted using time in study as the timescale including predictors selected by the lasso. Barlow weights with robust standard errors were used to account for the case-cohort design.^16,17^ Sub-cohort participants were weighted by the inverse of the sampling fraction and cases (COVID-19-related deaths) received a weight of 1 on the day they died and, unless they were also in the sub-cohort, a weight of zero prior to that. Royston-Parmar, Weibull and Generalised gamma models were also fitted. Results from these models are not shown: the Royston-Parmar models had very similar performance to the Cox models but the Weibull and Gamma models generally had poorer calibration. Note that these approach A models do not include any time-varying predictors or time-varying measures of prevalence of infection.

For approach B, data from all 73 sub-studies were stacked to form one analysis dataset, with a variable indicating the sub-study (k=1,2,…,73). A Poisson model for 28-day COVID-19 related death was fitted using Barlow weights with robust standard errors, incorporating predictors selected by the lasso and proxy measures of the burden of infection. Three sets of models were fitted: one for each of the three proxy measures. Weibull and logistic models were also fitted. Results from these models are not shown but performance was very similar to that of the Poisson.

For approach C, the series of 1-day studies were stacked to form one analysis dataset. A Poisson model was fitted to estimate the daily rate of COVID-19 related death using inverse sampling weights with robust standard errors, incorporating predictors selected by the lasso and proxy measures of the burden of infection. A similar Poisson model was fitted to estimate the daily rate of mortality due to non-COVID-19-related causes conditional on the same set of predictor variables, but without the measures of the burden of infection, weighted according to the inverse of the sampling fractions. Risk of 28-day COVID-19 related death was estimated by combining the estimates of daily survival from COVID-19 related death and other causes.

#### Missing data

The analysis was restricted to participants with recorded ethnicity data (which excluded 3,926,870 participants, approximately 26%). Participants with missing BMI were assumed non-obese and participants with no smoking information were assumed to be non-smokers, on the assumption that smoking and obesity, if present, are likely to be recorded. Patients with no serum creatinine measurement were included in the “no evidence of poor kidney function” group. Patients with diabetes but no glycosylated hemoglobin (HbA1c) measurement were included in a separate “diabetes, no HbA1c” category.

### Model validation

Three validation periods were considered, chosen to cover periods of higher and lower infection prevalence, within the period of time used for model development. Validation period 1 ran from 1^st^ March to 28^th^ March 2020, validation period 2 ran from 6^th^ April to 3^rd^ May 2020, validation period 3 ran from 12^th^ May to 8^th^ June 2020 (Figure 2). Internal validation was undertaken in each of these periods, assessing performance within the data the models were developed on. Geographical and temporal internal-external validation were also undertaken.^19^ For the geographical internal-external validation, a leave-one-out approach was used using the 7 regions of England, omitting all participants from one geographical region in turn, performing the model selection and fitting the model in the sub-sample excluding that region, and then using the fitted model, applied to the local prevalence measures in the omitted region for approaches B and C, to make predictions for the participants in the omitted region. This was repeated for each of seven regions. For the temporal internal-external validation, the data were split into two time-periods: 1^st^ March 2020 until 11^th^ May and 12^th^ May until 8^th^ June 2020. For each validation period, all eligible participants who remained alive at the start of the period were included in the validation. Risks of 28-day COVID-19 related death were predicted using each model, using predictors and proxy measures of infection prevalence (where used) from the start of the validation period. Model performance was reported by comparing the observed outcome, 28-day COVID-19 related death, to the predicted risk.

**Figure 2.**
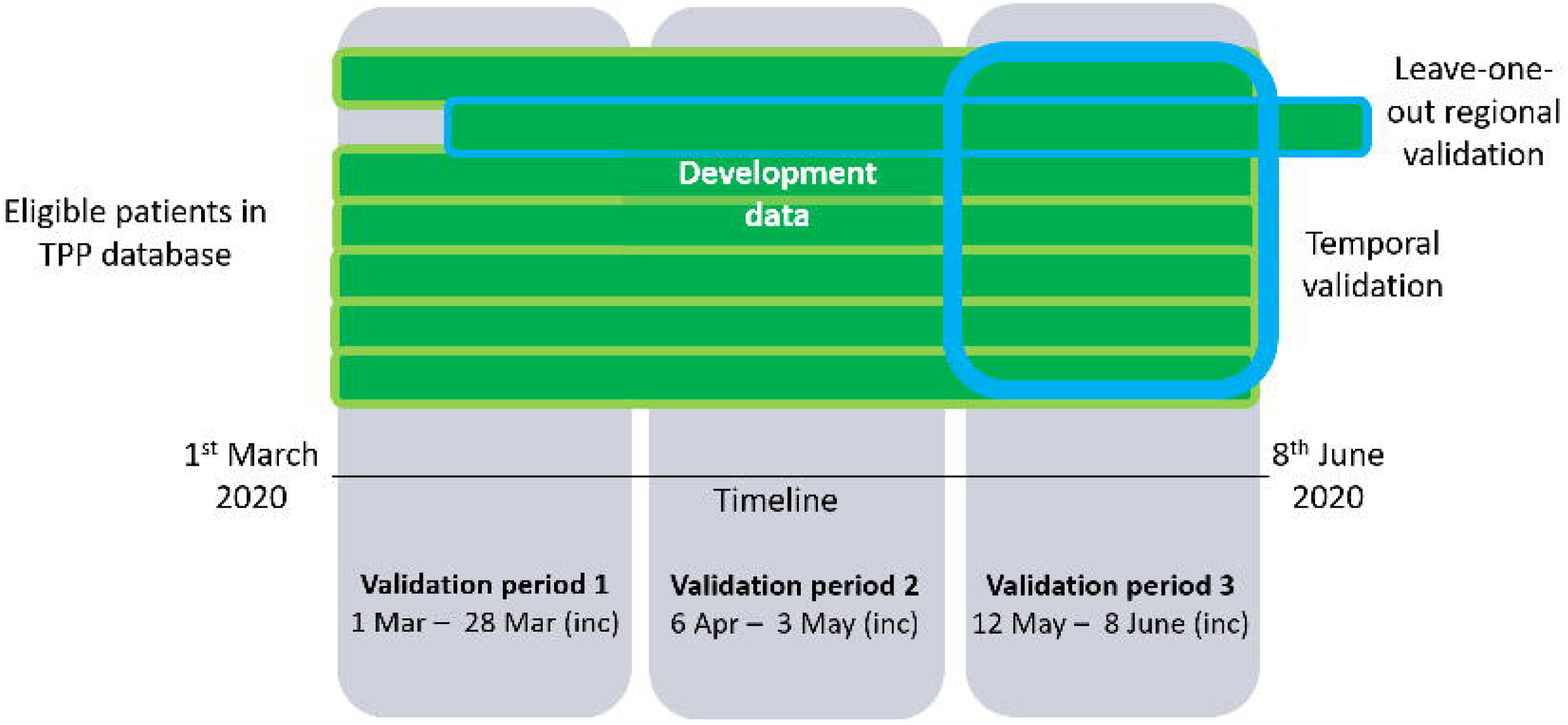
Schematic showing the internal and internal-external validation undertaken

**Figure 3.**
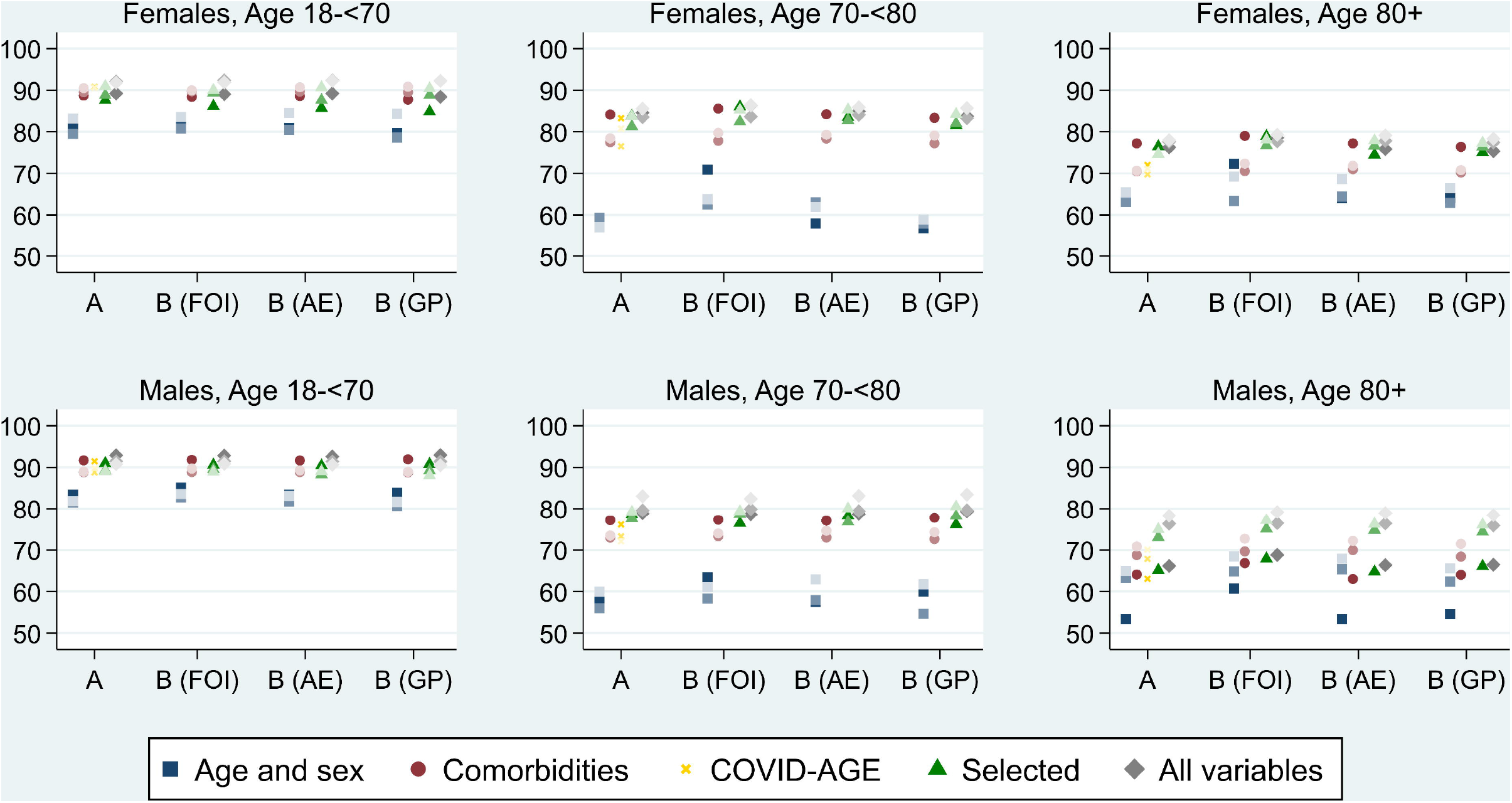
C-statistics for a range of models using modelling approach A and B, with the latter incorporating modelled estimates of the COVID-19 infection prevalence (ME), rates of A&E COVID-19 attendance (AE) and suspected COVID-19 rates in primary care (GP). Models shown are: Age and sex only, model including comorbidities via the total number only, the COVID-AGE tool, the main selected model, and a richer model including all variables. The three points within each graph, for each approach, show the three validation periods, differentiated by depth of colour (deepest = validation period 1, lightest = validation period 3).

Discrimination – the ability to distinguish between cases (COVID-19 related deaths) and non-cases – was assessed by Harrell’s C-statistic, expressed as a percentage.^20,21^ The C-statistic estimates the probability that, if a case and non-case are randomly selected, the predicted risk will be higher for the case than the non-case. A completely uninformative prediction would have a C-statistic of 50% and a perfect prediction would have a C-statistic of 100%. Calibration – the agreement between observed outcomes and predictions – was assessed by (i) comparing mean predicted risk with mean observed risk, (ii) by estimating the calibration intercept and slope, with 95% confidence intervals, to assess whether models over-or under-estimate risk.^21,22^ The calibration intercept estimates the mean difference between observed and predicted risks, with intercepts less than zero indicating over-prediction on average and intercepts greater than zero indicating under-prediction. The calibration slope estimates the relationship between the log-odds of the observed risk and the log-odds of the predicted risk, with slopes greater than one indicating that risk is under-estimated in high risk and over-estimated in low risk participants; the opposite holds for slopes of less than one. We reported model performance overall and within sex and broad age-group (18-<70, 70-<80 and 80+). Insufficient events occurred in the youngest age-group to split further.

A pre-specified analysis, described in the protocol, was to compare performance of the lasso-selected models to models including fewer potential predictors. We considered: (i) a model including only age-group (10 categories: <40, 40-49, 50-59, 60-64, 65-69, 70-74, 75-79, 80-84, 85-89, 90+), sex and their interaction; (ii) number of comorbidities (0, 1, 2, 3+), sex, age-group, with all three-way interactions between these variables, ethnicity and a rural/urban indicator; (iii) the COVID-AGE risk tool (10^th^ update).^6,7^ For approach B models, the proxy measures of burden of infection were additionally included.

The comorbidities included in the “number of comorbidities” variable for model (ii) were: respiratory disease, cystic fibrosis, severe asthma (with OCS use), chronic cardiac disease, atrial fibrillation, deep vein thrombosis or pulmonary embolism, surgery for arterial disease, diabetes, cancer diagnosed in the last year, haematological malignancy diagnosed within 5 years, liver disease, stroke, dementia, neurological disease, poor kidney function (eGFR <60 mL/min/1.73m^2^), dialysis, organ transplant, asplenia, condition inducing immunosuppression, HIV, and either obese or underweight.

For the COVID-AGE risk tool, we could not reliably distinguish Type I from Type II diabetes so included all diabetes as Type II. We did not have an indicator of heart failure but included chronic cardiac disease. COVID-AGE is primarily a risk stratification tool, rather than intended to estimate absolute risks, so we did not estimate calibration measures for this tool.

We also fitted a richer model including all candidate predictors without any variable selection, with all interactions with age (linear) and sex and additional spline terms for age.

### Software and reproducibility

Data management was performed using Python and Google BigQuery, with analysis carried out using Stata 16.1 / Python. All of the code used for data management and analyses is openly shared online for review and re-use under MIT open license (https://github.com/opensafely/risk-prediction-research).

### Patient and Public Involvement

We have developed a publicly available website https://opensafely.org/ through which we invite any patient or member of the public to contact us regarding this study or the broader OpenSAFELY project.

## RESULTS

Table 1 described the base cohort used in these analyses, comprising almost 12 million patients of whom 7,999 experienced a COVID-19 related death. The case-cohort sample used for approach A included all 7,999 COVID-19 related deaths and a sub-cohort of 319,917, which contained 683 of the COVID-19 related deaths. The stacked landmark studies included all 7,999 COVID-19 deaths and 330,132 participants.

**Table 1.** Description of cohort used for model development

|  | N (%) | COVID-19 related deaths (%) |
| --- | --- | --- |
| Total | 11,972,947 (100.0) | 7,999 (0.07) |
| *Age-group:* |  |  |
| 18-39 | 4,275,852 (35.7) | 52 (0.00) |
| 40-49 | 2,022,527 (16.9) | 141 (0.01) |
| 50-59 | 2,040,181 (17.0) | 437 (0.02) |
| 60-69 | 1,635,143 (13.7) | 938 (0.06) |
| 70-79 | 1,319,367 (11.0) | 2,025 (0.15) |
| 80+ | 6,798,77 (5.7) | 4,406 (0.65) |
| *Sex:* |  |  |
| Female | 6,232,725 (52.1) | 3,315 (0.05) |
| Male | 5,740,222 (47.9) | 4,684 (0.08) |
| *BMI:* |  |  |
| Underweight (<18.5) | 249,294 (2.1) | 306 (0.12) |
| No evidence of underweight/obesity | 3,982,133 (33.3) | 2,480 (0.06) |
| Obese I (30-34.9) | 3,690,583 (30.8) | 2,430 (0.07) |
| Obese II (35-39.9) | 1,794,812 (15.0) | 1,462 (0.08) |
| Obese III (40+) | 678,109 (5.7) | 658 (0.10) |
| Missing | 1,242,341 (10.4) | 340 (0.03) |
| *Smoking:* |  |  |
| Never smoker | 5,540,732 (46.3) | 2,499 (0.05) |
| Former smoker | 3,921,016 (32.7) | 4,745 (0.12) |
| Current smoker | 2,253,231 (18.8) | 737 (0.03) |
| Missing | 257,968 (2.2) | 18 (0.01) |
| *Ethnicity:* |  |  |
| White | 10,184,871 (85.1) | 6,952 (0.07) |
| Indian | 405,477 (3.4) | 299 (0.07) |
| Pakistani | 262,882 (2.2) | 161 (0.06) |
| Bangladeshi/Other Asian | 262,882 (2.2) | 161 (0.06) |
| African/Other black | 280,466 (2.3) | 173 (0.06) |
| Caribbean | 80,863 (0.7) | 124 (0.15) |
| Chinese | 103,423 (0.9) | 20 (0.02) |
| Mixed/Other | 392,097 (3.3) | 147 (0.04) |
| *Deprivation:* |  |  |
| IMD 1 (least deprived) | 2,315,449 (19.3) | 1,255 (0.05) |
| IMD 2 | 2,375,974 (19.8) | 1,443 (0.06) |
| IMD 3 | 2,398,815 (20.0) | 1,537 (0.06) |
| IMD 4 | 2,489,997 (20.8) | 1,791 (0.07) |
| IMD 5 (most deprived) | 2,392,712 (20.0) | 1,973 (0.08) |
| *Location:* |  |  |
| Urban | 9,595,617 (80.1) | 6,775 (0.07) |
| Rural | 2,377,330 (19.9) | 1,224 (0.05) |
| *Region:* |  |  |
| East | 2,730,203 (22.8) | 1773 (0.06) |
| London | 1,018,332 (8.5) | 755 (0.07) |
| Midlands | 2,673,963 (22.3) | 2005 (0.07) |
| North East and Yorkshire | 2,242,375 (18.7) | 1679 (0.07) |
| North West | 1,053,537 (8.8) | 905 (0.09) |
| South East | 744,930 (6.2) | 309 (0.04) |
| South West | 1,509,607 (12.6) | 573 (0.04) |
| *Blood pressure:* |  |  |
| Normal | 2,797,632 (23.4) | 1,725 (0.06) |
| Elevated | 1,757,455 (14.7) | 1,406 (0.08) |
| High, stage I | 3,899,203 (32.6) | 2,454 (0.06) |
| High, stage II | 2,492,161 (20.8) | 2,389 (0.10) |
| Missing | 1,026,496 (8.6) | 25 (0.00) |
| Diagnosed hypertension | 2,448,605 (20.5) | 5,332 (0.22) |
| **Comorbidities** |  |  |
| ***Respiratory*** |  |  |
| Asthma, no OCS use | 1,829,710 (15.3) | 1,008 (0.06) |
| Asthma, with OCS use | 112,407 (0.9) | 248 (0.22) |
| Respiratory disease | 495,699 (4.1) | 1,809 (0.36) |
| Cystic Fibrosis or other conditions | 3,167 (0.0) | 4 (0.13) |
| ***Cardiovascular*** |  |  |
| Cardiac disease | 783,896 (6.5) | 3,008 (0.38) |
| Atrial Fibrillation | 430,798 (3.6) | 1,808 (0.42) |
| DVT/PE | 249,969 (2.1) | 773 (0.31) |
| PAD or lower limb amputation | 41,362 (0.3) | 220 (0.53) |
| Diabetes, controlled | 738,369 (6.2) | 1,908 (0.26) |
| Diabetes, uncontrolled | 346,726 (2.9) | 1,054 (0.30) |
| Diabetes, status unknown | 133,387 (1.1) | 306 (0.23) |
| ***Neurological*** |  |  |
| Stroke | 240,401 (2.0) | 1,287 (0.54) |
| Vascular dementia | 22,792 (0.2) | 489 (2.15) |
| Other neurological condition | 114,431 (1.0) | 480 (0.42) |
| ***Cancer (non-haematological)*** |  |  |
| Diagnosed <1 year ago | 54,290 (0.5) | 216 (0.40) |
| Diagnosed 2-5 years ago | 157,859 (1.3) | 358 (0.23) |
| Diagnosed 5+ years ago | 362,457 (3.0) | 879 (0.24) |
| ***Haematological cancer*** |  |  |
| Diagnosed <1 year ago | 6,151 (0.1) | 47 (0.76) |
| Diagnosed 2-5 years ago | 18,722 (0.2) | 95 (0.51) |
| Diagnosed 5+ years ago | 42,492 (0.4) | 116 (0.27) |
| ***Kidney and liver*** |  |  |
| Reduced kidney function<br>(eGFR in range 30-<60 mL/min/1.73m<sup>2</sup>) | 607,308 (5.1) | 2,793 (0.46) |
| Very reduced kidney function<br>(eGFR <30 mL/min/1.73m<sup>2</sup>) | 58,081 (0.5) | 694 (1.19) |
| Dialysis | 8,871 (0.1) | 115 (1.30) |
| Liver disease | 74,193 (0.6) | 189 (0.25) |
| Organ transplant | 11,349 (0.1) | 50 (0.44) |
| ***Immunosuppression*** |  |  |
| Spleen | 19,815 (0.2) | 29 (0.15) |
| RA/SLE/Psoriasis | 609,421 (5.1) | 733 (0.12) |
| Immunosuppression | 13,091 (0.1) | 28 (0.21) |
| HIV | 23,078 (0.2) | 17 (0.07) |
| Inflammatory bowel disease | 152,080 (1.3) | 169 (0.11) |
| **Other** |  |  |
| Fracture (in >65 year old in last 2 years) | 55,952 (0.5) | 443 (0.79) |
| Learning disability | 158,350 (1.3) | 161 (0.10) |
| Serious mental illness | 150,928 (1.3) | 254 (0.17) |

Table 2 shows measures of model performance in predicting 28-day risk in the three 28-day validation periods. Estimated model coefficients are provided in the appendix. For all models in all validation periods, the C-statistic was high (92-94%), indicating excellent ability to distinguish between cases and non-cases. For approach A, the Cox model, which did not explicitly model the prevalence of infection, the mean predicted and observed risks were very similar in the first validation period, but different in the second and third. In validation period 2, the mean observed risk was ten times higher than in validation period 1, but the mean predicted risk using approach A was (by design) the same for all three periods. The poor calibration in validation periods 2 and 3 was also reflected in high calibration intercepts.

**Table 2.** Measures of model performance in predicting 28-day risk of COVID-19 mortality

| Approach | Measures of infection prevalence included | Model | Validation period | C-statistic (%) | Observed mean risk (%) | Predicted mean risk (%) | Calibration |  |
| --- | --- | --- | --- | --- | --- | --- | --- | --- |
|  |  |  |  |  |  |  | Intercept (95% CI) | Slope (95% CI) |
| A | None | Cox | 1 | 92.4 | 0.0038 | 0.0038 | 0.00 (-0.10, 0.09) | 0.95 (0.86, 1.05) |
|  |  |  | 2 | 93.4 | 0.0374 | 0.0038 | 2.30 (2.27, 2.32) | 1.02 (0.99, 1.05) |
|  |  |  | 3 | 94.1 | 0.0104 | 0.0037 | 1.03 (0.97, 1.08) | 1.05 (1.00, 1.11) |
| B | Modelled estimates | Poisson Approach B | 1 | 92.5 | 0.0038 | 0.0044 | -0.15 (-0.24, -0.06) | 0.93 (0.84, 1.02) |
|  |  |  | 2 | 93.7 | 0.0374 | 0.0354 | 0.06 (0.03, 0.09) | 1.00 (0.97, 1.03) |
|  |  |  | 3 | 94.4 | 0.0104 | 0.0128 | -0.20 (-0.26, -0.15) | 1.03 (0.98, 1.09) |
| B | A&E COVID-19 attendances | Poisson | 1 | 92.1 | 0.0038 | 0.0145 | -1.34 (-1.43, -1.25) | 0.92 (0.83, 1.02) |
|  |  |  | 2 | 93.3 | 0.0374 | 0.0420 | -0.12 (-0.15, -0.09) | 0.99 (0.96, 1.02) |
|  |  |  | 3 | 94.3 | 0.0104 | 0.0197 | -0.64 (-0.69, -0.58) | 1.05 (0.99, 1.10) |
| B | Suspected COVID-19 in primary care | Poisson | 1 | 92.1 | 0.0038 | 0.0085 | -0.80 (-0.89, -0.71) | 0.90 (0.81, 1.00) |
|  |  |  | 2 | 93.5 | 0.0374 | 0.0378 | -0.01 (-0.04, 0.02) | 1.00 (0.97, 1.03) |
|  |  |  | 3 | 94.2 | 0.0104 | 0.0156 | -0.41 (-0.46, -0.35) | 1.04 (0.98, 1.09) |

For the approach B model incorporating the modelled estimates of infection prevalence, the mean and observed risks were very similar in all three validation periods. The calibration intercept was slightly less than zero in validation periods 1 and 3, indicating slight over-estimation on average, with calibration slopes close to one, indicating reasonable calibration. Replacing the modelled estimates by either the rate of A&E COVID-19 attendances or the rate of suspected COVID-19 cases in primary care resulted in poorer calibration, particularly in the first validation period which had a very low infection prevalence. All approach B models had worse calibration than the approach A Cox model in validation period 1, the data on which the models are fitted, but considerably better calibration than the approach A model in the other two validation periods. All approach B models had very high discrimination, with C-statistics (92-94%).

Approach C models performed similarly to approach B when using the actual measures of infection prevalence through the 28 day period. However, poor estimation of the measure using only data available at day 0 of the period led, in some cases, to large under- and over-estimation of risks (Table A3).

The internal-external validation showed that the performance of the approach A Cox model was insensitive to the removal of a region or period of time (Appendix Figure A1). Approach B models were more sensitive to geographical or temporal omissions.

Among the oldest age group (80+ years), discrimination was substantially lower than in the younger age-groups, reflecting the substantial discrimination that comes simply from age. Generally, discrimination was a little lower among males. The comparison of the discrimination by models that considered few versus many potential predictors, found that a simple model including only age and sex still provided fairly good discrimination (∼80%) among the 18-<70 age-group, but was substantially worse for the two older age-groups. In all cases, the model including the number of comorbidities, rather than a large number of indicators of individual comorbidities, performed almost as well as the models that considered many more predictors. COVID-AGE, which we applied only for approach A, performed similarly to the most complex models used.

## DISCUSSION

We have developed and validated a risk prediction model for COVID-19 mortality in the general population with very high discrimination which can be applied within primary care electronic health record (EHR) systems without requiring linkage to external data, allowing accurate prediction of relative risk. In addition, we have developed a second set of models that enable estimation of absolute risk by incorporating estimates of the prevalence of COVID-19 infection. Incorporating modelled estimates led to the best model performance, but this may be impractical for most situations due to the complex modelling required to obtain these estimates. Incorporating the rate of suspected COVID-19 cases in local primary care practices led to reasonable model performance; this rate could be quantified within EHR systems in an automated manner, providing timely absolute risk estimates as the pandemic evolves. Finally, our third set of models using daily landmarking allow estimation and comparison of absolute COVID-19 mortality risks under various scenarios (e.g. drastically increasing infection rates versus gradual decrease).

We found that the existing COVID-AGE tool^6,7^ had very high discrimination, similar to the best performing models considered, suggesting this provides a reliable ranking of COVID-19 mortality risk. Interestingly, very simple models including only age, sex, ethnicity, a rural indicator, and a count of total comorbidities led to models with very good discrimination. When focusing on specific patient groups with higher morbidity, more complex models may provide useful additional discrimination, but in many cases much simpler models are able to discriminate well.

In extending our knowledge of which approaches to use in predicting related COVID-19 death from a general population to inform future policies, this study has three major strengths. First, this is the first risk prediction algorithm that incorporates time-varying estimates of infection burden to provide accurate estimates of the absolute risk of COVID-19 related death. Second, the study uses data from a large representative sample of the general population. Third, we consider three different designs, several alternative estimation approaches, and contrast different ways of selecting predictors.

This study also has important weaknesses. First, the COVID-19 pandemic is evolving rapidly. We will re-validate and, if necessary, re-calibrate our models using data from the more recent wave, particularly in light of evidence suggesting differences in mortality with the new variant. Further, as the vaccines are rolled out, risk in the population will additionally depend on proportions of different patient groups vaccinated; this could be incorporated into subsequent iterations of these models. We have not yet externally validated these models. However, our internal-external validation suggests little over-optimism in the measures of model performance.

Electronic health record data are not collected for research, so information on certain characteristics can be incomplete or absent. For example, ascertainment of HIV and cystic fibrosis is likely to be poor and pregnancy cannot easily be identified, limiting the ability to distinguish risks between these groups.

Our approach to missing data reflected the way in which these models might be used in practice if applied within electronic health record systems. For example, patients with no BMI measurement would be assumed to have normal BMI. Our measures of model performance reflect performance under this implementation. The exception is ethnicity – given the strong relationships previously observed between ethnicity and COVID-19 outcomes we chose to restrict our sample to those with recorded ethnicity. In previous work, model estimates from similar models were similar in ethnicity-complete-case samples and following multiple imputation.^10^

We have explored risk prediction for COVID-19 death in the general population. In some circumstances, the risk of COVID-19 death given infection with the virus might be of interest, and this could not be estimated with the present data in the absence of representative COVID-19 testing data. However, risk within the general population is more useful for various policy decisions, such as vaccine prioritisation.

We did not attempt to compare the performance of our risk prediction models to QCOVID,^6^ because our database does not include all data required to implement that algorithm at present. In comparison to QCOVID, our algorithm used a smaller set of potential predictors. For example, we did not include homelessness as a predictor. Further, we restricted our sample to adults living in the community, so our risk predictions are not valid for those living in residential care. QCOVID reported a C-statistic of 93%; our models ranged from 92 to 94%.

Our results have a number of implications for policy makers and GPs advising patients. Most of the discriminating power in each model evaluated here was driven by simple features such as age, sex and a count of co-morbidities. Complex risk prediction models driven by multiple variables from diverse sources can be difficult, slow, and expensive to implement in routine care: the cost, opportunity costs and complexity of such implementations may not be warranted. The fact that very simple models produce very high discrimination also suggests that policies targeting population level reduction of COVID-19 mortality risk may not need to distinguish between all comorbidities in detail. For certain policy decisions, such as vaccine prioritisation, this simplicity would provide a great advantage, with simple eligibility criteria resulting in faster programme rollout and delivery of vaccines.

When absolute risk estimates are obtained from algorithms that do not explicitly model the prevalence of infection it is important to consider the context that data were collected in, in order to avoid misleading interpretations of absolute risk estimates. Our validated landmarking models can avoid such problems by providing absolute risk estimates which explicitly account for rapidly changing infection levels.

Risk scores in COVID-19 need to be a dynamic and open process undergoing regular transparent re-calibration rather than a single “one-off” digital commodity. We have developed a completely open source, transparent and freely available set of risk prediction models for COVID-19 mortality. Our approach accounts for rapid changes in the estimates of infection burden, due to daily changes in for example the number of people vaccinated, and therefore provide accurate estimates of the absolute risk of COVID-19 related deaths to inform the ongoing policy debate.

## Data Availability

NHS England is the data controller; TPP is the data processor; and the key researchers on OpenSAFELY are acting on behalf of NHS England. This implementation of OpenSAFELY is hosted within the TPP environment which is accredited to the ISO 27001 information security standard and is NHS IG Toolkit compliant; patient data has been pseudonymised for analysis and linkage using industry standard cryptographic hashing techniques; all pseudonymised datasets transmitted for linkage onto OpenSAFELY are encrypted; access to the platform is via a virtual private network (VPN) connection, restricted to a small group of researchers; the researchers hold contracts with NHS England and only access the platform to initiate database queries and statistical models; all database activity is logged; only aggregate statistical outputs leave the platform environment following best practice for anonymisation of results such as statistical disclosure control for low cell counts. Detailed information on all code lists is openly shared at https://codelists.opensafely.org/ for inspection and reuse.

## DECLARATIONS

## Acknowledgements

This work uses data provided by patients and collected by the NHS as part of their care and support. We are very grateful for all the support received from the TPP Technical Operations team throughout this work, and for generous assistance from the information governance and database teams at NHS England / NHSX. We thank Professor Rafael Perera, Nuffield Department of Primary Care Health Science, for useful comments.

## Data Sharing

All data were linked, stored and analysed securely within the OpenSAFELY platform https://opensafely.org/. Data include pseudonymized data such as coded diagnoses, medications and physiological parameters. No free text data are included. Detailed pseudonymised patient data is potentially re-identifiable and therefore not shared. We rapidly delivered the OpenSAFELY data analysis platform without prior funding to deliver timely analyses on urgent research questions in the context of the global Covid-19 health emergency: now that the platform is established we are developing a formal process for external users to request access in collaboration with NHS England; details of this process will be published shortly on https://opensafely.org/.

## Transparency declaration

BG affirms that this manuscript is an honest, accurate and transparent account of the study being reported; that no important aspects of the study have been omitted; and that any discrepancies from the study as planned have been explained.

## Information governance and ethical approval

NHS England is the data controller; TPP is the data processor; and the key researchers on OpenSAFELY are acting on behalf of NHS England. This implementation of OpenSAFELY is hosted within the TPP environment which is accredited to the ISO 27001 information security standard and is NHS IG Toolkit compliant;^18,19^ patient data has been pseudonymised for analysis and linkage using industry standard cryptographic hashing techniques; all pseudonymised datasets transmitted for linkage onto OpenSAFELY are encrypted; access to the platform is via a virtual private network (VPN) connection, restricted to a small group of researchers; the researchers hold contracts with NHS England and only access the platform to initiate database queries and statistical models; all database activity is logged; only aggregate statistical outputs leave the platform environment following best practice for anonymisation of results such as statistical disclosure control for low cell counts.20 The OpenSAFELY research platform adheres to the obligations of the UK General Data Protection Regulation (GDPR) and the Data Protection Act 2018. In March 2020, the Secretary of State for Health and Social Care used powers under the UK Health Service (Control of Patient Information) Regulations 2002 (COPI) to require organisations to process confidential patient information for the purposes of protecting public health, providing healthcare services to the public and monitoring and managing the COVID-19 outbreak and incidents of exposure; this sets aside the requirement for patient consent.^21^ Taken together, these provide the legal bases to link patient datasets on the OpenSAFELY platform. GP practices, from which the primary care data are obtained, are required to share relevant health information to support the public health response to the pandemic, and have been informed of the OpenSAFELY analytics platform.

This study was approved by the Health Research Authority (REC reference 20/LO/0651) and by the LSHTM Ethics Board (reference 21863).

## Contributorship

B.G. conceived the platform and the approach; B.G. and L.S. led the project overall and are guarantors; S.B. led the software; E.J.W, J.T. and K.B. led the statistical analysis; C.E.M. and A.J.W. led on codelists and implementation; and A.M. led on information governance. Contributions are as follows: data curation, C.B., J.P., J.C., S.H., S.B., D.E., P.I. and C.E.M.; analysis, E.J.W., K.B., A.J.W. and C.E.M.; funding acquisition, B.G. and L.S.; information governance, A.M., B.G., C.B. and J.P.; methodology, E.J.W., K.B., A.J.W., B.G., L.S., C.B., J.P., J.C., S.H., S.B., D.E., P.I., C.E.M., R.G., D.H., R.K. K. D-O, E.S. and R.P.; disease category conceptualization and codelists, C.E.M., A.J.W., P.I., S.B., D.E., C.B., J.C., J.P., S.H., H.J.C., K.B., S.B., A.M., B.M., L.T., I.J.D., H.I.M., R.M. and H.F.; ethics approval, H.J.C., E.J.W., L.S. and B.G.; project administration, C.E.M., H.J.C., C.B., S.B., A.M., L.S. and B.G.; resources, B.G., L.S. and F.H.; software, S.B., D.E., P.I., A.J.W., C.E.M., C.B., F.H., J.C. and S.H.; supervision, B.G., L.S. and S.B.; writing (original draft), E.J.W., J.T. All authors were involved in design and conceptual development and reviewed and approved the final manuscript.

## REFERENCES

1. WHO. WHO Director-General’s opening remarks at the media briefing on COVID-19: 11 March 2020. who.int. Published 2020. https://web.archive.org/web/20200502133342/https://www.who.int/dg/speeches/detail/who-director-general-s-opening-remarks-at-the-media-briefing-on-covid-1911-march-2020

2. WHO. COVID-19 situation reports. who.int. Published January 2021. https://www.who.int/publications/m/item/weekly-epidemiological-update27-january-2021

3. UK Government. Number of coronavirus (COVID-19) cases and risk in the UK. gov.uk. Published 2020. https://coronavirus.data.gov.uk/?_ga=2.27729908.99180100.1612170744-430880118.1599131957&_gac=1.45721558.1612020033.CjwKCAiApNSABhAlEiwANuR9YC39VvGHldtXUCLD4ys8_tznWOYGzhcio-LmcQxV8vX9iwYwPiA9sBoCZTMQAvD_BwE

4. UK Government. COVID-19: comparison of geographic allocation of cases in England by lower tier local authority. gov.uk. Published 2020. https://www.gov.uk/government/publications/covid-19-comparison-of-geographic-allocation-of-cases-in-england-by-lower-tier-local-authority

5. Wynants L, Van Calster B, Collins GS, et al. Prediction models for diagnosis and prognosis of covid-19: systematic review and critical appraisal. BMJ. 369:m1328.

6. Covid-19 Medical Risk Assessment (10th update). alama.org.uk. https://alama.org.uk/covid-19-medical-risk-assessment/

7. Coggon D, Croft P, Cullinan P, Williams A. Assessment of workers’ personal vulnerability to covid-19 using ‘covid-age.’ Occup Med. 2020;70(7):461–464. doi:10.1093/occmed/kqaa150

8. Clift AK, Coupland CA, Keogh R, et al. QCOVID a living risk prediction algorithm for risk of hospitalisation and mortality from COVID-19 in adults: national derivation and validation cohort study. BMJ.

9. Wolff R, Moons KGM, Riley R, et al. PROBAST: A Tool to Assess the Risk of Bias and Applicability of Prediction Model Studies. Ann Intern Med. 2019;170(1).

10. Williamson EJ, Walker AJ, Bhaskaran K, et al. OpenSAFELY: factors associated with COVID-19 death in 17 million patients. Nature. Published online July 8, 2020. doi:10.1038/s41586-020-2521-4

11. Williamson E, Tazare J, Bhaskaran K, et al. Study protocol: Comparison of different risk prediction modelling approaches for COVID-19 related death using the OpenSAFELY platform [version 1; peer review: 1 approved]. Wellcome Open Res. 2020;5(243). doi:10.12688/wellcomeopenres.16353.1

12. Collins G, Reitsma, JB, Altman D, Moons KGM. Transparent reporting of a multivariable prediction model for individual prognosis or diagnosis (TRIPOD): the TRIPOD statement. BMJ. 2015;350.

13. Davies NG, Kucharski AJ, Eggo RM, et al. Effects of non-pharmaceutical interventions on COVID-19 cases, deaths, and demand for hospital services in the UK: a modelling study. Lancet Public Health. 2020;5(7):e375–e385. doi:10.1016/S2468-2667(20)30133-X

14. van Houwelingen HC, Putter H. Dynamic predicting by landmarking as an alternative for multi-state modeling: an application to acute lymphoid leukemia data. Lifetime Data Anal. 2008;14(4):447–463. doi:10.1007/s10985-008-9099-8

15. Lau B, Cole SR, Gange SJ. Competing Risk Regression Models for Epidemiologic Data. Am J Epidemiol. 2009;170(2):244–256. doi:https://doi.org/10.1093/aje/kwp107

16. Barlow W, Ichikawa L, Rosner D, Izumi S. Analysis of case-cohort designs. J Clin Epidemiol. 1999;52(11):1165–1172.

17. Onland-Moret NC, van der A DL, van der Schouw YT, et al. Analysis of case-cohort data: A comparison of different methods. J Clin Epidemiol. 2007;60(4):350–355. doi:10.1016/j.jclinepi.2006.06.022

18. Pavlou M, Ambler G, Seaman S, De Iorio M, Omar RZ. Review and evaluation of penalised regression methods for risk prediction in low-dimensional data with few events. Stat Med. 2016;35(7):1159–1177. doi:10.1002/sim.6782

19. Steyerberg EW, Harrell FE Jr. Prediction models need appropriate internal, internal-external, and external validation. J Clin Epidemiol. 2016;69:245–247. doi:10.1016/j.jclinepi.2015.04.005

20. Newson RB. Comparing the predictive powers of survival models using Harrell’s C or Somers’ D. Stata J. 2010;10(3):339–358.

21. Steyerberg EW, Vickers AJ, Cook NR, et al. Assessing the performance of prediction models: a framework for traditional and novel measures. Epidemiol Camb Mass. 2010;21(1):128–138. doi:10.1097/EDE.0b013e3181c30fb2

22. Royston P. Tools for Checking Calibration of a Cox Model in External Validation: Approach Based on Individual Event Probabilities. Stata J. 2014;14(4):738–755. doi:10.1177/1536867X1401400403

