## Supplementary Appendix for "Predicting COVID-19 related death using the OpenSAFELY platform"

28 January 2021

The OpenSAFELY Collaborative, Elizabeth J Williamson*, John Tazare*, Krishnan Bhaskaran, Helen I McDonald, Alex J Walker, Laurie Tomlinson, Kevin Wing, Sebastian Bacon, Chris Bates, Helen J Curtis, Harriet Forbes, Caroline Minassian, Caroline E Morton, Emily Nightingale, Amir Mehrkar, Dave Evans, Brian D Nicholson, Dave Leon, Peter Inglesby, Brian MacKenna, Nicholas G Davies, Jonathan Cockburn, Will Hulme, Jess Morley, Ian Douglas, Christopher T Rentsch, Rohini Mathur, Angel Wong, Anna Schultze, Richard Croker, John Parry, Frank Hester, Sam Harper, Richard Grieve, David A Harrison, Ewout W. Steyerberg, Rosalind M Eggo, Karla Diaz-Ordaz, Ruth Keogh, Stephen JW Evans, Liam Smeeth†, Ben Goldacre† (ORCID: 0000-0002-5127-4728).

† Joint principal investigators; * joint first authors

A1. Potential predictors

All codelists used to define predictors are listed in Table A1 below.

Age (continuous); sex (male or female); e[thnicity](https://codelists.opensafely.org/codelist/opensafely/ethnicity/) (8 category: White, Indian, Pakistani, Bangladeshi/Other Asian, African/Other black, Caribbean, Chinese, Mixed/Other); deprivation (quintile of the index of multiple deprivation (IMD) derived from the patient’s postcode at lower super output area level); the number of adults living in the household (continuous) and whether or not children aged up to 12 years are living in the household (yes/no); the region (seven regions of England: South West, South East, London, East, Midlands, North West, and North East, Yorkshire and the Humber) and whether the individual lives in a rural or urban area were included.

Obesity was grouped using categories derived from the World Health Organisation classification of Body Mass Index (BMI; kg/m^2^): underweight <18.5 kg/m^2^; obese I 30-34.9; obese II 35-39.9; obese III 40+; or no evidence of obesity or being underweight, with BMI ascertained from weight measurements within the last 10 years, restricted to those taken when the patient was over 16 years old. Smoking status was grouped into evidence of current smoking in the last 18 months, former and never smokers.

Comorbidities were defined through combinations of clinical measurements, prescriptions, and recorded diagnoses. Blood pressure (in a measurement taken in the last 18 months) was grouped into: high II (systolic blood pressure (SBP) >= 140 mmHg or diastolic blood pressure (DBP) >= 90 mmHg), high I (SBP 130-<140 or DBP 80-<90), elevated (SPB 120-<130 and DBP <80) or normal (SBP<120 and DBP < 80). Where the two measures (SBP and DBP) differed, the higher (worse) class was taken. Diagnosed hypertension; [chronic cardiac disease](https://codelists.opensafely.org/codelist/opensafely/chronic-cardiac-disease/) including chronic heart failure, ischaemic heart disease, and severe valve or congenital heart disease likely to require lifelong follow up; atrial fibrillation; surgery for peripheral arterial disease or lower limb amputation; prior deep vein thrombosis or pulmonary embolism; [diabetes](https://codelists.opensafely.org/codelist/opensafely/diabetes/) (additionally using HbA1c within last 15 months to determine level of HbA1c control, grouped into <58 mmol/mol (good control), >=58 mmol/mol (poor control) and no recent measure); stroke; [dementia](https://codelists.opensafely.org/codelist/opensafely/dementia/); and o[ther neurological conditions](https://codelists.opensafely.org/codelist/opensafely/other-neurological-conditions/) (motor neurone disease, myasthenia gravis, multiple sclerosis, Parkinson's disease, cerebral palsy, quadriplegia or hemiplegia, malignant primary brain tumour, and progressive cerebellar disease) were included.

Asthma (grouped by use of oral corticosteroids as an indication of severity, with 2 or more prescriptions in the last year taken to indicate severe asthma); cystic fibrosis and associated diseases such as primary ciliary dyskinesia; and other r[espiratory disease](https://codelists.opensafely.org/codelist/opensafely/chronic-respiratory-disease/); haematological malignancies (considered separately from other cancers to reflect the immunosuppression associated with haematological malignancies and their treatment) and non-haematological malignancy, each grouped according to time since diagnosis (<1 year, 2-<5 years, 5+years); l[iver disease](https://codelists.opensafely.org/codelist/opensafely/chronic-liver-disease/); solid organ transplant; dialysis, for patients who have not since had a kidney transplant; and kidney function were included. Kidney function was ascertained from the most recent serum creatinine measurement taken in the last 5 years excluding the most recent fortnight, where available, converted into estimated glomerular filtration rate (eGFR) using the Chronic Kidney Disease Epidemiology Collaboration (CKD-EPI) equation), with reduced kidney function grouped into no evidence of kidney impairment (no creatinine measurement or eGFR>=60 mL/min/1.73m^2^), stage 3 (eGFR in range 30-<60 mL/min/1.73m^2^) and stage 4-5 (<30 mL/min/1.73m^2^). Patients with a history of kidney dialysis or kidney transplant were included in the category representing stage 4-5.

Rheumatoid Arthritis (RA), Systemic Lupus Erythematosus (SLE) or psoriasis; asplenia (splenectomy or a spleen dysfunction, including sickle cell disease); other immunosuppressive conditions including a condition inducing permanent immunodeficiency ever diagnosed, or aplastic anaemia or temporary immunodeficiency recorded within the last year; inflammatory bowel disease; HIV; learning disability, including Down’s syndrome; serious mental illness; and fragility fracture in the last two years for patients aged 65 or above were also included.

Table A1. Codelists used to define candidate predictors and suspected COVID-19 (one of the proxy measures of infection burden)

| Variable | Notes | Codelist |
| --- | --- | --- |
| Suspected COVID-19 |  | <https://codelists.opensafely.org/codelist/opensafely/covid-identification-in-primary-care-suspected-covid-suspected-codes/2020-07-16/> |
| Ethnicity | 7 categories, obtained from 16 (White, Caribbean, Chinese, Indian/Pakistani, Mixed/Other, Bangladeshi/Other Asian, African/Other Black) | [https://codelists.opensafely.org/codelist/opensafely/ethnicity](https://codelists.opensafely.org/codelist/opensafely/ethnicity/) |
| Diagnosed hypertension |  | [https://codelists.opensafely.org/codelist/opensafely/hypertension](https://codelists.opensafely.org/codelist/opensafely/hypertension/) |
| C[hronic cardiac disease](https://codelists.opensafely.org/codelist/opensafely/chronic-cardiac-disease/) |  | <https://codelists.opensafely.org/codelist/opensafely/chronic-cardiac-disease/> |
| Atrial Fibrillation |  | <https://codelists.opensafely.org/codelist/opensafely/atrial-fibrillation-or-flutter/2020-07-30/> |
| Surgery for Peripheral Arterial Disease | Combined with lower limb amputation to form a peripheral arterial disease variable | <https://codelists.opensafely.org/codelist/opensafely/surgery-for-peripheral-artery-disease/2020-09-16/> |
| Lower limb amputation | Combined with surgery for peripheral arterial disease to form a peripheral arterial disease variable | <https://codelists.opensafely.org/codelist/opensafely/amputation/2020-09-21/> |
| Prior deep vein thrombosis / pulmonary embolism |  | <https://codelists.opensafely.org/codelist/opensafely/venous-thromboembolic-disease/2020-09-14/> |
| D[iabetes](https://codelists.opensafely.org/codelist/opensafely/diabetes/) | Combined with HbA1c measure within 18 months to determine level of control | <https://codelists.opensafely.org/codelist/opensafely/diabetes/> |
| S[troke](https://codelists.opensafely.org/codelist/opensafely/stroke-updated/) |  | <https://codelists.opensafely.org/codelist/opensafely/stroke/> |
| Dementia |  | <https://codelists.opensafely.org/codelist/opensafely/dementia/> |
| O[ther neurological conditions](https://codelists.opensafely.org/codelist/opensafely/other-neurological-conditions/) |  | <https://codelists.opensafely.org/codelist/opensafely/other-neurological-conditions/> |
| Asthma | Combined with OCS prescriptions in past year to determine severity | <https://codelists.opensafely.org/codelist/opensafely/asthma-diagnosis/> |
| Cystic Fibrosis and associated conditions |  | <https://codelists.opensafely.org/codelist/opensafely/cystic-fibrosis/2020-07-20/> |
| R[espiratory disease other than asthma](https://codelists.opensafely.org/codelist/opensafely/chronic-respiratory-disease/) or cystic fibrosis |  | <https://codelists.opensafely.org/codelist/opensafely/other-chronic-respiratory-disease/2020-07-20/> |
| Non-haematological cancer | Grouped by time since diagnosis (<1 year, 2-<5 years, 5+years) | <https://codelists.opensafely.org/codelist/opensafely/cancer-excluding-lung-and-haematological/> |
| Haematological cancer | Grouped by time since diagnosis (<1 year, 2-<5 years, 5+years) | <https://codelists.opensafely.org/codelist/opensafely/haematological-cancer/> |
| Lung cancer | Combined with other non-haematological cancer | <https://codelists.opensafely.org/codelist/opensafely/lung-cancer/> |
| Chronic l[iver disease](https://codelists.opensafely.org/codelist/opensafely/chronic-liver-disease/) |  | <https://codelists.opensafely.org/codelist/opensafely/chronic-liver-disease/> |
| Kidney dialysis | Used if no kidney transplant since most recent dialysis | <https://codelists.opensafely.org/codelist/opensafely/dialysis/2020-07-16/> |
| Kidney transplant | Combined with non-kidney transplant for transplant indicator. Also used to determine which of dialysis/transplant is most recent. | <https://codelists.opensafely.org/codelist/opensafely/kidney-transplant/2020-07-15/> |
| Organ transplant (other than kidney) | Combined with kidney transplant for transplant indicator. | <https://codelists.opensafely.org/codelist/opensafely/other-organ-transplant/2020-07-15/> |
| A[splenia](https://codelists.opensafely.org/codelist/opensafely/asplenia/) or dysplenia |  | <https://codelists.opensafely.org/codelist/opensafely/asplenia/> <https://codelists.opensafely.org/codelist/opensafely/sickle-cell-disease/> |
| Autoimmune diseases ([rheumatoid arthritis, lupus, psoriasis](https://codelists.opensafely.org/codelist/opensafely/ra-sle-psoriasis/)) |  | <https://codelists.opensafely.org/codelist/opensafely/ra-sle-psoriasis/> |
| HIV |  | [https://codelists.opensafely.org/codelist/opensafely/hiv/2020-07-13/](https://codelists.opensafely.org/codelist/opensafely/hiv/2020-07-13/#full-list) |
| Other immunosuppressive condition | Temporary and aplastic anaemia within last year; permanent ever. | <https://codelists.opensafely.org/codelist/opensafely/permanent-immunosuppresion/>  <https://codelists.opensafely.org/codelist/opensafely/aplastic-anaemia/>  <https://codelists.opensafely.org/codelist/opensafely/temporary-immunosuppresion/> |
| Inflammatory bowel disease |  | <https://codelists.opensafely.org/codelist/opensafely/inflammatory-bowel-disease/2020-04-07/> |
| Learning disability, including Down’s syndrome |  | <https://codelists.opensafely.org/codelist/opensafely/intellectual-disability-including-downs-syndrome/2020-08-27/> |
| Serious mental illness |  | <https://codelists.opensafely.org/codelist/opensafely/psychosis-schizophrenia-bipolar-affective-disease/2020-07-09/> |
| Fragility fracture |  | <https://codelists.opensafely.org/codelist/opensafely/fragility/2020-09-14/> |

A2. Estimated regression coefficients

Table A2. Estimated Hazard Ratios from a Cox model, approach A

| Characteristic | Hazard Ratio (95% CI) |
| --- | --- |
| Age | 7.56 (7.16, 7.98) |
| Male | 2.13 (1.97, 2.29) |
| Urban | 1.52 (1.42, 1.62) |
| IMD 1 | 0.79 (0.74, 0.84) |
| IMD 5 | 1.33 (1.22, 1.45) |
| Ethnicity: White | 0.58 (0.54, 0.63) |
| BMI: Underweight | 1.53 (1.33, 1.75) |
| BMI: Normal/overweight | 0.81 (0.77, 0.86) |
| Diabetes: None | 0.68 (0.64, 0.72) |
| Diabetes: Control unknown | 1.22 (1.07, 1.39) |
| No stroke | 0.71 (0.64, 0.79) |
| No dementia | 0.36 (0.30, 0.42) |
| No other neurological | 0.14 (0.10, 0.18) |
| Asthma: With OCS | 1.23 (1.06, 1.43) |
| No respiratory | 0.35 (0.28, 0.42) |
| Cancer (exc. haematological): Last year | 1.63 (1.34, 2.00) |
| Cancer (haematological): Never | 0.44 (0.37, 0.53) |
| No liver disease | 0.27 (0.17, 0.41) |
| Renal impairment: None | 0.70 (0.65, 0.76) |
| Renal impairment: Stage 4/5 | 10.08 (7.24, 14.03) |
| No immunosuppression | 0.47 (0.30, 0.74) |
| No serious mental illness | 0.41 (0.36, 0.48) |
| Interactions with age: |  |
| BMI: Obese II (Per unit age increase) | 1.14 (1.06, 1.21) |
| BMI: Obese III (Per unit age increase) | 0.94 (0.79, 1.12) |
| Current smoker (Per unit age increase) | 0.97 (0.91, 1.03) |
| Hypertension (Per unit age increase) | 0.98 (0.95, 1.01) |
| Diabetes: Uncontrolled (Per unit age increase) | 0.99 (0.92, 1.07) |
| Other neurological (Per unit age increase) | 0.51 (0.43, 0.61) |
| Respiratory (Per unit age increase) | 0.70 (0.62, 0.78) |
| Cancer (haem): Last year (Per unit age increase) | 1.07 (0.78, 1.46) |
| Liver disease (Per unit age increase) | 0.62 (0.46, 0.85) |
| Dialysis (Per unit age increase) | 1.45 (1.17, 1.80) |
| Renal impairment: Stage 4/5 (Per unit age increase) | 0.37 (0.31, 0.44) |
| Asplenia (Per unit age increase) | 1.01 (0.77, 1.33) |
| IBD (Per unit age increase) | 1.11 (1.00, 1.23) |
| Fracture (Per unit age increase) | 1.25 (1.16, 1.34) |
| Interactions with sex: |  |
| (In males) IMD 5 | 0.97 (0.86, 1.08) |
| (In females) BMI: Obese III | 1.83 (1.36, 2.48) |
| (In males) Diabetes: Uncontrolled | 1.39 (1.19, 1.62) |
| (In females) Cardiac disease | 1.30 (1.19, 1.41) |
| (In females) Atrial Fibrillation | 1.22 (1.11, 1.34) |
| (In males) Surgery for PAD | 1.72 (1.41, 2.09) |
| (In males) Stroke | 1.05 (0.91, 1.21) |
| (In males) Dementia | 1.22 (0.97, 1.54) |
| (In females) Asthma: Without OCS | 1.07 (0.96, 1.18) |
| (In females) Respiratory | 1.26 (1.12, 1.42) |
| (In females) Cancer (exc. haematological): Last year | 2.01 (1.49, 2.72) |
| (In males) Cancer (exc. haematological): 2-5 years ago | 1.25 (1.09, 1.44) |
| (In males) Cancer (exc. haematological): 5+ years ago | 1.06 (0.96, 1.17) |
| (In females) Cancer (haematological): Last year | 1.28 (0.54, 3.02) |
| (In males) Cancer (haematological): 5+ years ago | 0.60 (0.44, 0.82) |
| (In females) Liver disease | 1.27 (0.88, 1.82) |
| (In females) Dialysis | 1.61 (0.98, 2.63) |
| (In females) Renal impairment: Stage 4/5 | 1.25 (1.02, 1.54) |
| (In males) Renal impairment: Stage 3a/3b | 0.96 (0.86, 1.07) |
| (In males) RA/SLE/Psoriasis | 1.12 (1.00, 1.26) |
| (In males) Fracture | 1.65 (1.32, 2.06) |

Table A3: Estimated Incidence Rate Ratios from a Poisson model, approach B, incorporating the modelled estimates as a proxy for the infection prevalence

| Characteristic | Incidence Rate Ratio (95% CI) |
| --- | --- |
| Age | 7.66 (7.24,8.10) |
| Male | 2.13 (1.97, 2.31) |
| Log of modelled estimate | 1.91 (1.85,1.97) |
| Qd(ME)* | 7.98 (3.79,16.83) |
| Urban | 1.41 (1.27, 1.56) |
| IMD 1 | 0.47 (0.39, 0.56) |
| Ethnicity: Mixed | 1.23 (1.02, 1.47) |
| BMI: Obese II | 2.05 (1.69, 2.49) |
| Diabetes: None | 0.67 (0.63, 0.71) |
| Diabetes: Uncontrolled | 1.52 (1.37, 1.69) |
| No cardiac disease | 0.79 (0.75, 0.84) |
| No DVT/PE | 0.64 (0.59, 0.70) |
| No other neurological | 0.44 (0.38, 0.51) |
| No respiratory | 0.59 (0.54, 0.64) |
| Cancer (exc. haematological): Last year | 10.47 (7.22, 15.18) |
| No organ transplant | 0.47 (0.31, 0.72) |
| Renal impairment: None | 0.67 (0.61, 0.73) |
| Renal impairment: Stage 4/5 | 1.89 (1.61, 2.21) |
| No intellectual disability | 0.23 (0.19, 0.28) |
| No fracture | 0.49 (0.42, 0.56) |
| Number in household (spline term 2) | 2.53 (1.59,4.02) |
| Number in household (spline term 3) | 0.19 (0.05,0.68) |
| Interactions with age: |  |
| IMD 1 (Per unit age increase) | 1.38 (1.24, 1.53) |
| IMD 5 (Per unit age increase) | 1.13 (1.08, 1.19) |
| Ethnicity: Indian (Per unit age increase) | 0.98 (0.90, 1.08) |
| Ethnicity: Pakistani (Per unit age increase) | 0.88 (0.77, 1.00) |
| BMI: Obese II (Per unit age increase) | 0.79 (0.69, 0.91) |
| BMI: Obese III (Per unit age increase) | 1.06 (0.88, 1.26) |
| BP Normal (Per unit age increase) | 1.23 (1.18, 1.28) |
| BP Stage I (Per unit age increase) | 0.95 (0.92, 0.98) |
| Atrial Fibrillation (Per unit age increase) | 1.09 (1.04, 1.14) |
| Surgery for PAD (Per unit age increase) | 1.30 (1.18, 1.42) |
| Stroke (Per unit age increase) | 1.08 (1.03, 1.15) |
| Dementia (Per unit age increase) | 1.48 (1.35, 1.62) |
| Asthma: Without OCS (Per unit age increase) | 0.97 (0.92, 1.01) |
| Asthma: With OCS (Per unit age increase) | 0.98 (0.85, 1.12) |
| Cancer (exc. haematological): Last year (Per unit age increase) | 0.35 (0.28, 0.43) |
| Cancer (exc. haematological): 2-5 years ago (Per unit age increase) | 1.12 (1.05, 1.21) |
| Cancer (exc. haematological): 5+ years ago (Per unit age increase) | 1.01 (0.96, 1.05) |
| Cancer (haematological): 5+ years ago (Per unit age increase) | 1.20 (1.06, 1.35) |
| Dialysis (Per unit age increase) | 0.43 (0.29, 0.63) |
| Asplenia (Per unit age increase) | 1.02 (0.77, 1.35) |
| Serious mental illness (Per unit age increase) | 1.42 (1.19, 1.69) |
| Interactions with sex: |  |
| (In males) Rural | 0.87 (0.76, 1.00) |
| (In females) IMD 5 | 1.06 (0.94, 1.19) |
| (In females) BMI: Obese III | 1.68 (1.24, 2.29) |
| (In females) Diabetes: Uncontrolled | 1.01 (0.86, 1.19) |
| (In females) Atrial Fibrillation | 1.03 (0.90, 1.17) |
| (In females) Stroke | 1.18 (1.01, 1.39) |
| (In males) Dementia | 1.52 (1.18, 1.96) |
| (In females) Other neurological | 1.24 (0.97, 1.57) |
| (In females) Asthma: With OCS | 2.26 (1.70, 3.01) |
| (In females) Respiratory | 1.19 (1.05, 1.35) |
| (In females) Cancer (exc. haematological): Last year | 1.76 (1.29, 2.40) |
| (In males) Cancer (haematological): Last year | 3.26 (1.97, 5.41) |
| (In males) Cancer (haematological): 2-5 years ago | 2.61 (1.96, 3.48) |
| (In females) Dialysis | 14.60 (7.65, 27.89) |
| (In males) Dialysis | 7.24 (3.83, 13.68) |
| (In males) Renal impairment: Stage 3a/3b | 0.88 (0.79, 0.99) |
| (In males) Renal impairment: Stage 4/5 | 0.82 (0.66, 1.01) |
| (In females) IBD | 1.18 (0.91, 1.54) |
| (In females) Serious mental illness | 1.31 (0.91, 1.89) |
| (In males) Fracture | 1.36 (1.09, 1.70) |
| Constant | 0.01 (0.01,0.02) |

* Standardised quadratic coefficient obtained by fitting a quadratic model to the last three weeks of the measure, and dividing the linear coefficient by the constant coefficient.

A3. Approach C, daily landmarking: Measures of model performance

Table A4. Measures of model performance using approach C, daily landmarking. Proxy measures of the burden of infection are required through the 28 day validation period. These are obtained in three ways: (i) the actual measure as it evolves (which would not be available in practice), (ii) assuming the measure remains constant from the beginning of the validation period, and (iii) using a fractional polynomial model fitted to the prior 3 weeks of data.

| Burden of infection proxy measure used | Validation period | C-statistic | Observed mean risk (%) | Predicted mean risk (%) | Estimated calibration intercept  (95% CI) | Estimated calibration slope  (95% CI) |
| --- | --- | --- | --- | --- | --- | --- |
| **Actual measure** **obtained through 28 day validation period** | | | | | | |
| Modelled estimates | 1 | 92.6 | 0.0038 | 0.0064 | -0.52 (-0.62, -0.43) | 0.91 (0.81, 1.00) |
|  | 2 | 93.6 | 0.0374 | 0.0333 | 0.12 (0.09, 0.15) | 0.98 (0.96, 1.01) |
|  | 3 | 94.4 | 0.0104 | 0.0108 | -0.03 (-0.09, 0.02) | 1.03 (0.97, 1.08) |
| A&E COVID-19 attendance rates | 1 | 92.8 | 0.0038 | 0.0065 | -0.53 (-0.62, -0.44) | 0.95 (0.85, 1.04) |
|  | 2 | 93.2 | 0.0374 | 0.0347 | 0.08 (0.05, 0.10) | 0.98 (0.95, 1.01) |
|  | 3 | 94.2 | 0.0104 | 0.0113 | -0.08 (-0.13, -0.02) | 0.99 (0.93, 1.05) |
| Suspected COVID-19 case rates in primary care | 1 | 92.3 | 0.0038 | 0.0047 | -0.21 (-0.31, -0.12) | 0.93 (0.83, 1.02) |
|  | 2 | 93.3 | 0.0374 | 0.0328 | 0.13 (0.10, 0.16) | 0.98 (0.95, 1.01) |
|  | 3 | 94.0 | 0.0104 | 0.0142 | -0.31 (-0.37, -0.26) | 1.01 (0.96, 1.07) |
| **Assuming measure remains constant from day 0 of validation period** | | | | | | |
| Modelled estimates | 1 | 92.4 | 0.0038 | 0.0014 | 1.03 (0.94, 1.12) | 0.90 (0.81, 0.99) |
|  | 2 | 93.7 | 0.0374 | 0.0316 | 0.17 (0.14, 0.20) | 0.99 (0.96, 1.01) |
|  | 3 | 94.4 | 0.0104 | 0.0107 | -0.03 (-0.08, 0.03) | 1.02 (0.97, 1.08) |
| A&E COVID-19 attendance rates | 1 | 92.2 | 0.0038 | 0.0018 | 0.78 (0.68, 0.87) | 0.92 (0.83, 1.01) |
|  | 2 | 93.2 | 0.0374 | 0.0439 | -0.16 (-0.19, -0.13) | 0.98 (0.95, 1.01) |
|  | 3 | 94.2 | 0.0104 | 0.0160 | -0.43 (-0.49, -0.38) | 1.00 (0.95, 1.06) |
| Suspected COVID-19 case rates in primary care | 1 | 92.0 | 0.0038 | 0.0006 | 1.86 (1.77, 1.95) | 0.91 (0.81, 1.00) |
|  | 2 | 93.2 | 0.0374 | 0.0485 | -0.26 (-0.29, -0.23) | 0.98 (0.95, 1.00) |
|  | 3 | 94.0 | 0.0104 | 0.0177 | -0.53 (-0.59, -0.48) | 1.02 (0.97, 1.08) |
| **Predicted measure based on fractional polynomial model of previous 3 weeks of data** | | | | | | |
| Modelled estimates | 1 | 92.6 | 0.0038 | 0.0040 | -0.06 (-0.15, 0.03) | 0.91 (0.82, 1.00) |
|  | 2 | 63.2 | 0.0374 | 13.392 | * | * |
|  | 3 | 82.8 | 0.0104 | 3.3729 | * | * |
| A&E COVID-19 attendance rates | 1 | 92.2 | 0.0038 | 0.0018 | 0.78 (0.68, 0.87) | 0.92 (0.83, 1.01) |
|  | 2 | 92.1 | 0.0374 | 0.0392 | -0.05 (-0.08, -0.02) | 0.77 (0.74, 0.80) |
|  | 3 | 93.6 | 0.0104 | 0.02242 | -0.77 (-0.83, -0.71) | 0.94 (0.88, 0.99) |
| Suspected COVID-19 case rates in primary care | 1 | 91.9 | 0.0038 | 0.00315 | 0.19 (0.09, 0.28) | 0.90 (0.8, 0.99) |
|  | 2 | 90.1 | 0.0374 | 0.10474 | -1.06 (-1.09, -1.03) | 0.77 (0.74, 0.80) |
|  | 3 | 93.1 | 0.0104 | 0.00909 | 0.14 (0.08, 0.19) | 0.93 (0.87, 0.98) |

* Unable to be estimated (due to huge overestimation of risk overall)

Figure A1. Measures of model performance overall (Main), in the geographical internal-external validation (R_1_, …R_7_; omitting regions: East, London, Midlands, North East and Yorkshire, North West, South East and South West, respectively) and the temporal internal-external validation (R_-T_), in the three validation periods.

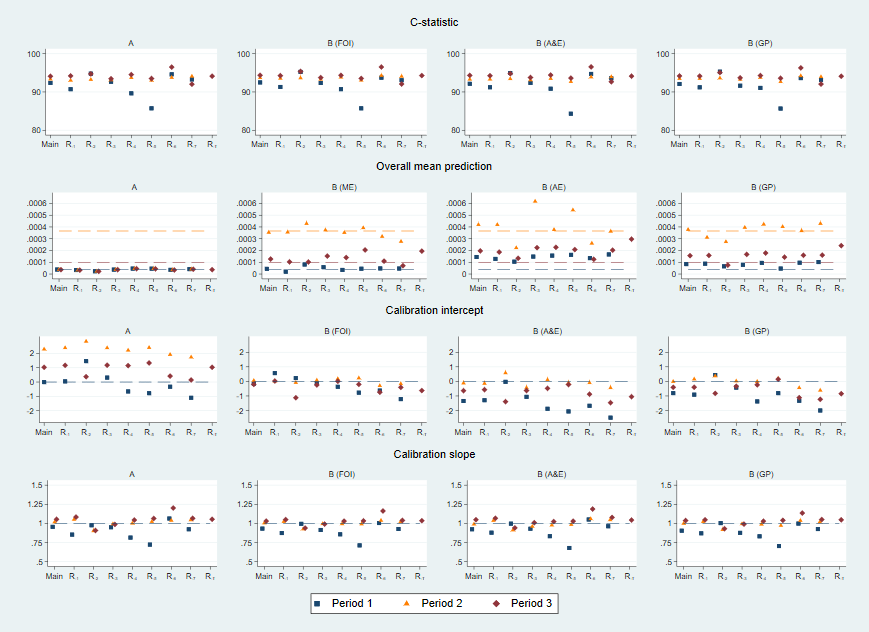
